# Soluble and cell-based markers of immune checkpoint inhibitor associated nephritis

**DOI:** 10.1101/2022.10.13.22280966

**Authors:** Meghan E. Sise, Qiyu Wang, Harish Seethapathy, Daiana Moreno, Destiny Harden, R. Neal Smith, Ivy A. Rosales, Robert B. Colvin, Sarah Chute, Lynn D. Cornell, Sandra Herrmann, Riley Fadden, Ryan J. Sullivan, Nancy Yang, Sara Barmettler, Alexandra Chloe Villani, Kerry Reynolds, Jocelyn Farmer

## Abstract

**Background:** Non-invasive biomarkers of immune checkpoint inhibitor-associated acute tubulointerstitial nephritis (ICI-nephritis) are urgently needed. Because ICIs block immune checkpoint pathways that include cytotoxic T lymphocyte antigen 4 (CTLA4), we hypothesized that biomarkers of immune dysregulation previously defined in patients with congenital CTLA4 deficiency, including elevated soluble interleukin-2 receptor alpha (sIL-2R) and flow cytometric cell-based markers of B and T cell dysregulation in peripheral blood may aide the diagnosis of ICI-nephritis.

**Methods:** A retrospective cohort of patients diagnosed with ICI-nephritis was compared to three prospectively enrolled control cohorts: ICI-treated controls without immune related adverse events, patients not on ICIs with hemodynamic acute kidney injury (hemodynamic AKI), and patients not on ICIs with biopsy proven acute interstitial nephritis from other causes (non-ICI-nephritis). sIL-2R level and flow cytometric parameters were compared between groups using Wilcoxon rank sum test or Kruskal-Wallis test. Receiver operating characteristic curves were generated to define the accuracy of sIL-2R and flow cytometric biomarkers in diagnosing ICI-nephritis. The downstream impact of T cell activation in the affected kidney was investigated using archived biopsy samples to evaluate the gene expression of *IL2RA*, IL-2 signaling, and T cell receptor signaling in patients with ICI-nephritis compared to other causes of drug-induced nephritis, acute tubular injury, and histologically normal controls.

**Results:** sIL-2R level in peripheral blood was significantly higher in patients with ICI-nephritis (N=24) (median 2.5-fold upper limit of normal [ULN], IQR 1.9-3.3), compared to ICI-treated controls (N=10) (median 0.8-fold ULN, IQR 0.5-0.9, *P*<0.001) and hemodynamic AKI controls (N=6) (median 0.9-fold-ULN, IQR 0.7-1.1, *P*=0.008). A sIL-2R cut-off point of 1.75-fold ULN was highly diagnostic of ICI-nephritis (AUC >96%) when compared to either ICI-treated or hemodynamic AKI controls. By peripheral blood flow cytometry analysis, lower absolute CD8+ T cells, CD45RA+CD8+ T cells, memory CD27+ B cells, and expansion of plasmablasts were prominent features of ICI-nephritis compared to ICI-treated controls. Gene expression for *IL2RA*, IL-2 signaling, and T cell receptor signaling in the kidney tissue with ICI-nephritis were significantly higher compared to controls.

**Conclusion:** Elevated sIL-2R level and flow cytometric markers of both B and T cell dysregulation may aid the diagnosis of ICI-nephritis.

**Key Messages:** *What is already known on this topic:* There are no non-invasive biomarkers of immune checkpoint inhibitor-associated nephritis (ICI-nephritis); kidney biopsy, the gold standard for diagnosing ICI-nephritis, can be challenging or even contraindicated given its periprocedural risk. There are mechanistic and clinicopathologic similarities between immune-related adverse events and congenital CTLA4 deficiency.

*What this study adds:* Established biomarkers of congenital CTLA4 deficiency, including elevated serum sIL-2R level and flow cytometric markers of both B and T cell dysregulation, are promising biomarkers for diagnosis of ICI-nephritis. These markers are not altered in patients treated with immune checkpoint inhibitors who are not experiencing immune-related adverse events.

*How this study might affect research, practice or policy:* Prospective study with longitudinal sIL-2R and peripheral flow cytometry measurements are needed to validate the result and may limit the need for invasive diagnosis of ICI-nephritis.

## Introduction

Immune checkpoint inhibitors (ICIs) significantly increase survival in patients with multiple cancer types and are changing the landscape of oncology care. To date, eight agents targeting the programmed cell death protein 1 (PD-1)/programmed cell death ligand 1 (PD-L1) signaling pathway or the cytotoxic T lymphocyte antigen 4 (CTLA4) signaling pathway have been approved by the United States Food and Drug Administration to treat over 20 cancer types (1). The latest estimates suggest that approximately 38% of all patients with cancer qualify for ICI treatment (2). Despite their success, treatment-induced inflammatory side effects termed “immune-related adverse events” (irAEs) affect 60-80% of patients who receive ICIs (3). irAEs are caused by T cell overactivation from disinhibition of the immune checkpoints, which under normal immune homeostasis function to maintain self-tolerance and prevent autoimmunity (3, 4). In the kidney, the most common irAE is acute tubulointerstitial nephritis (ICI-nephritis), which is estimated to affect 3-5% of patients receiving ICIs (5, 6).

Acute kidney injury (AKI) is common in patients with cancer, affecting almost 25% of ICI-treated patients within the first year of treatment (7-9). The etiology of AKI in ICI-treated patients not only includes ICI-nephritis but also other causes such as acute tubular injury from sepsis and nephrotoxins, pre-renal azotemia from volume depletion, obstruction, and others (2). Obtaining kidney biopsy tissue for histological examination remains the gold standard for ICI-nephritis diagnosis, but this can be challenging in patients with cancer due to concerns about bleeding risk, infection risk, and/or patient preference (10-12). It is difficult to identify ICI-nephritis without a biopsy as clinical features, laboratory testing, urinalysis findings, and conventional imaging do not reliably distinguish ICI-nephritis from other common causes of AKI (13-15). Yet, important treatment decisions that can affect both kidney function and cancer outcomes depend on accurate diagnosis of ICI-nephritis. Given these diagnostic challenges, non-invasive biomarkers are needed.

Because ICIs block immune checkpoint pathways that include CTLA4, we hypothesized that knowledge gained from patients with congenital CTLA4 deficiency could be applied to the understanding of immune pathways mediating ICI-nephritis. CTLA4 deficiency is a rare inborn error of immunity due to germline loss-of-function mutations in *CTLA4* or its regulators (e.g., *LRBA* and *DEF6*). CTLA4 deficiency causes constitutive and pathologic T cell hyperactivation due to decreased CTLA4 levels and/or function in T regulatory cells. T cell hyperactivation drives end-organ auto-inflammatory disease, which is analogous to the irAEs triggered by ICIs. Detailed immunophenotyping in CTLA4 deficiency has demonstrated that loss of naïve T cells, as well as atypical activation and dysregulated maturation of the B cell compartment are cellular markers of disease (16, 17). Patients with CTLA4 deficiency also have increased levels of cytokines correlating with T cell activation, and soluble interleukin-2 receptor alpha (sIL-2R, aka soluble CD25), specifically, has been utilized both to risk stratify for end-organ autoimmune disease and to assess therapeutic response to immunomodulators (18, 19).

We hypothesized that peripheral blood biomarkers characteristic of CTLA4 deficiency, including an increased sIL-2R level and cell-based markers of T and B cell dysregulation measured by flow cytometry, would be found in the blood of patients with ICI-nephritis compared to ICI-treated controls. Additionally, we evaluated gene expression of *IL2RA*, the IL-2 signaling pathway, and the T cell receptor signaling pathway in kidney biopsies of patients with ICI-nephritis compared to controls to validate the downstream impact of these pathways in the kidney.

## Methods

### Study design and patient population

We retrospectively reviewed patients who were referred to an Onconephrology practice for evaluation of AKI after ICI treatment between September 2019 and February 2022. Cases of ICI-nephritis were confirmed by meeting the following definition: 1) 1.5-fold rise in serum creatinine from pre-ICI baseline creatinine within 3 months of having received an ICI and 2) kidney biopsy demonstrating acute interstitial nephritis, or in patients who did not undergo kidney biopsy, ICI-nephritis was clinically adjudicated by the treating nephrologist and oncologist after exclusion of alternate causes of AKI, which led to temporary discontinuation of ICI or corticosteroid use. Two nephrologists retrospectively reviewed and unanimously agreed that the case was ICI-nephritis. Cases that did not meet the above criteria for ICI-nephritis were categorized as possible ICI-nephritis and were not included in the generation of receiver operating curve (ROC).

We prospectively enrolled three control cohorts: ICI-treated controls, hemodynamic AKI controls, and non-ICI-nephritis controls. The first control cohort included patients who were receiving ICIs for treatment of cancer and had normal kidney function, no history of irAEs, and no concurrent or recent corticosteroid use (ICI-treated controls). The second control cohort included non-ICI-treated patients who were hospitalized and were experiencing hemodynamic AKI (hemodynamic AKI controls). The nephrologist-adjudicated etiology of AKI for hemodynamic AKI controls is shown in **Supplemental Table 1**. Finally, we enrolled a control cohort of non-ICI-treated patients with biopsy-proven acute tubulointerstitial nephritis from other causes (non-ICI-nephritis controls) (**Supplemental Table 2**).

Both cases and controls were excluded if they had an active infection at the time of sampling or if they received >2.5 mg/day of prednisone equivalent or any other immunosuppressive medication within 2 weeks of the sample collection. Additionally, patients with known diagnosis of hematological malignancy (e.g., chronic lymphocytic leukemia) were excluded in analysis of flow cytometry given significant alteration of lymphocyte markers due to underlying disease, but they remained in the analysis of sIL-2R.

This protocol was approved by the IRB at Mass General Brigham. The IRB waived the need for signed informed consent for retrospective data collection. Control patients signed informed consent for prospective sample collection.

### Blood sample processing

sIL-2R level and/or peripheral blood flow cytometry was performed on fresh blood samples from cases and controls prior to administration of corticosteroid therapy. sIL-2R was performed on a fresh serum sample. During the study period, sIL-2R was performed by either Quest (reference range 532-1891 pg/mL) or ARUP laboratories (reference range 175.3-858.2 pg/mL). Given that these two assays differed in median and range, results of sIL-2R were standardized to the fold-change from upper limit of normal (ULN) for each test, which was used for analysis rather than absolute values. All flow cytometry was performed on fresh samples (same-day processing) in the CLIA-certified Pathology Department at the Massachusetts General Hospital using two panels: 1) a combined panel of T cell markers (CD3, CD4, CD8, CD45RA, CD45RO) and B cell markers (CD19, CD27, IgM, IgD), and 2) an activated B cell/plasmablast subspecialty panel (plasmablasts defined by CD19+CD20-CD27+CD38+).

### Statistical analysis

Continuous variables are presented as median with interquartile range or geometric mean with geometric standard deviation factor as indicated. Categorical variables are presented as counts and/or percentages as indicated. Receiver operating characteristic (ROC) curves were generated for sIL-2R by comparing fold ULN sIL-2R level between ICI-nephritis cases and ICI-treated controls, as well as fold ULN sIL-2R level between ICI-nephritis cases and hemodynamic AKI controls. ROC curves were generated for single and/or composite flow cytometry data by comparing ICI-nephritis cases to ICI-treated controls. Wilcoxon rank sum test was used for two group comparisons of continuous variables and Kruskal-Wallis test was used for multiple group comparisons (≥3) of continuous variables. The optimal cut-offs were chosen based on maximizing specificity. All analyses were performed in Prism 9.

### Nanostring RNA assay and analysis

Routine formalin fixed, paraffin embedded (FFPE) biopsy blocks were retrieved from archived samples at Massachusetts General Hospital or Mayo clinic, separated from the aforementioned clinical cohorts. All biopsies had been obtained as a part of routine care and had sufficient remaining tissue after completion of diagnostic studies. The pathologic groups included acute interstitial nephritis due to ICI (N=22), drug-induced acute interstitial nephritis (N=19), acute tubular injury (N=9), and controls from histologically normal renal biopsies (N=11). Five or six consecutive 20-μm curls cut from each FFPE block of kidney tissue were immediately transferred to sterile microcentrifuge tubes and stored at room temperature. Deparaffinization and RNA extraction were performed using Quick-RNA FFPE Miniprep (Zymo Research, Irvine, CA, USA). RNA concentration and purity were measured with a Nano-Drop 2000 spectrophotometer (Thermo Fisher Scientific, Waltham, MA, USA). Gene expression of the FFPE tissue-derived RNA isolates was quantified using the nCounter MAX System (NanoString Technologies, Seattle, WA, USA). We used the Banff Human Organ Transplant (B-HOT) 770-gene panel for hybridization (NanoString Technologies).(20) This panel was selected because of the pathologic similarities of acute interstitial nephritis to acute cellular rejection, and because it is enriched for immune cell genes. QC assessment and normalization were performed as described.(21) The NanoString Advanced Analysis software 2.0 was used to define IL-2 signaling and T cell receptor pathway scores using the first principal component of the gene sets shown in **Supplemental Table 3** (22). Diagnostic groups were compared using Kruskal-Wallis test.

## Results

We identified 29 patients with suspected ICI-nephritis who had either sIL-2R or flow cytometry performed as a part of their evaluation of AKI (**Figure 1**). After adjudication, 24 were deemed to have ICI-nephritis (7 were biopsy proven, 17 clinically diagnosed), and 5 had “possible ICI-nephritis.” Only one patient in the ICI-nephritis group received chronic corticosteroid therapy (2.5mg daily) at baseline for treatment of polymyalgia rheumatica. After diagnosis and sample collection, 20 of the 24 patients (83%) with ICI-nephritis were subsequently treated with corticosteroids, and the remaining four were treated with supportive therapy alone (holding ICI and any potential nephritis-triggering medications). There were 10 ICI-treated controls with normal kidney function, 6 patients with hemodynamic AKI, and 5 patients with non-ICI-nephritis (**Figure 1**). Baseline characteristics of the cohorts are shown in **Table 1**.

**Table 1.**
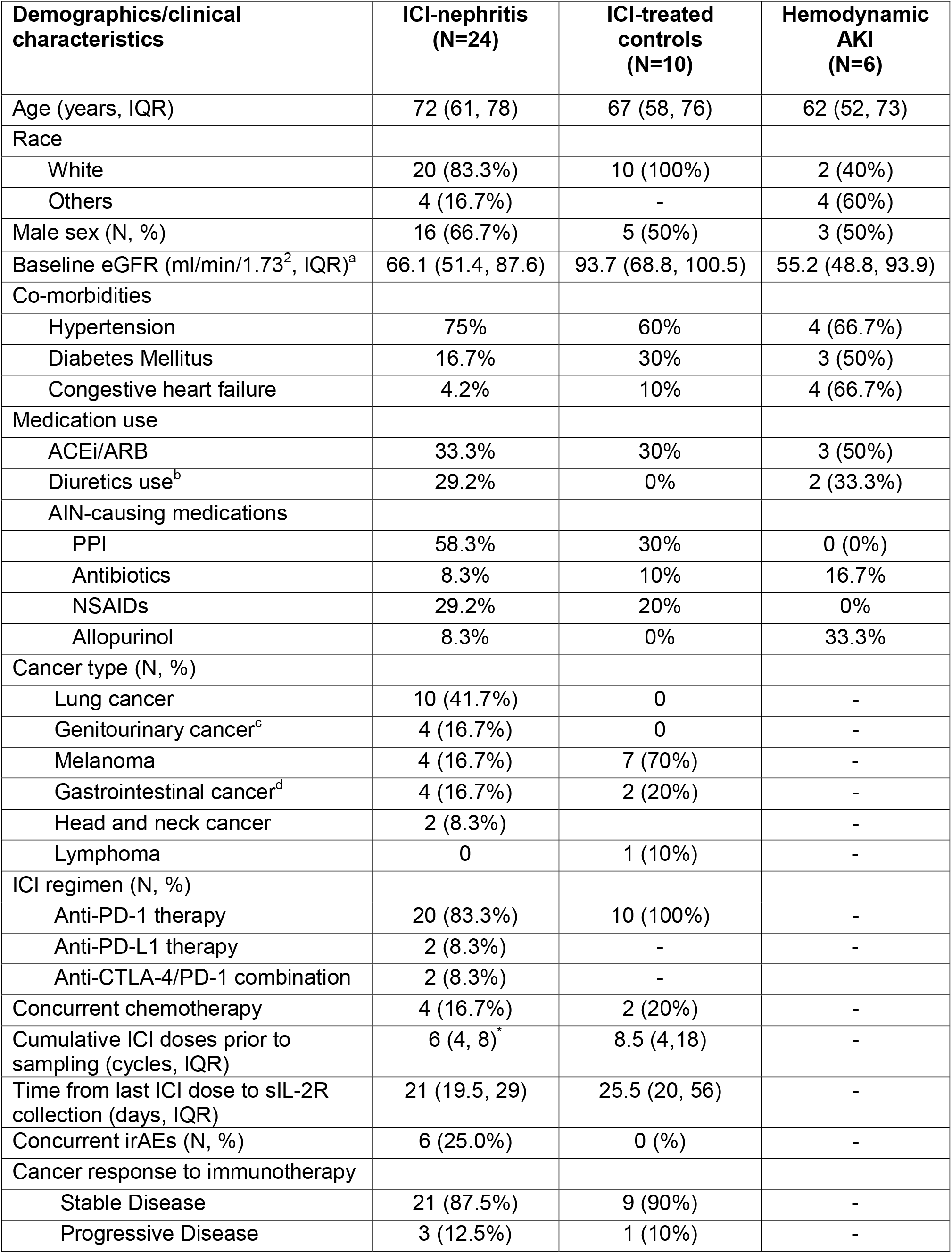
Demographics and baseline clinical characteristics of confirmed ICI-nephritis, ICI-treated controls, and hemodynamic AKI controls. a. Baseline eGFR was defined as the closest eGFR within 14 days prior to ICI initiation in ICI-nephritis group, closest eGFR within 14 days prior to sIL-2R level in ICI-treated controls, and closest available eGFR prior to AKI onset in the hemodynamic AKI group. Three patients in the hemodynamic AKI group did not have creatinine available within 90 days prior to AKI onset, and their baseline eGFR level was imputed from the nadir creatinine during hospitalization. b. Includes loop, thiazide-like, and potassium-sparing diuretics. c. Includes renal cell carcinoma, urothelial/transitional cell carcinoma. d. Includes gastric carcinoma, esophageal carcinoma, and pancreatic carcinoma. *one patient did not have known cumulative ICI doses (received from outside hospital) prior to sIL-2R sampling at our institution. Abbreviations: IQR=interquartile range; ACEi/ARB = angiotensin converting enzyme inhibitor/angiotensin receptor blockade; AIN=acute interstitial nephritis; PPI = proton pump inhibitor; NSAIDs = nonsteroidal anti-inflammatory drugs; anti-CTLA-4 = anti-cytotoxic T-lymphocyte-associated protein 4; anti-PD1= anti-programmed cell death protein1; anti-PD-L1=anti-programmed cell death protein1 ligand 1; irAEs=immune related adverse events.

| **Demographics/clinical characteristics** | **ICI-nephritis (N=24)** | **ICI-treated controls (N=10)** | **Hemodynamic AKI (N=6)** |
| --- | --- | --- | --- |
| Age (years, IQR) | 72 (61, 78) | 67 (58, 76) | 62 (52, 73) |
| Race |  |  |  |
| White | 20 (83.3%) | 10 (100%) | 2 (40%) |
| Others | 4 (16.7%) | - | 4 (60%) |
| Male sex (N, %) | 16 (66.7%) | 5 (50%) | 3 (50%) |
| Baseline eGFR (ml/min/1.73<sup>2</sup>, IQR)<sup>a</sup> | 66.1 (51.4, 87.6) | 93.7 (68.8, 100.5) | 55.2 (48.8, 93.9) |
| Co-morbidities |  |  |  |
| Hypertension | 75% | 60% | 4 (66.7%) |
| Diabetes Mellitus | 16.7% | 30% | 3 (50%) |
| Congestive heart failure | 4.2% | 10% | 4 (66.7%) |
| Medication use |  |  |  |
| ACEi/ARB | 33.3% | 30% | 3 (50%) |
| Diuretics use<sup>b</sup> | 29.2% | 0% | 2 (33.3%) |
| AIN-causing medications |  |  |  |
| PPI | 58.3% | 30% | 0 (0%) |
| Antibiotics | 8.3% | 10% | 16.7% |
| NSAIDs | 29.2% | 20% | 0% |
| Allopurinol | 8.3% | 0% | 33.3% |
| Cancer type (N, %) |  |  |  |
| Lung cancer | 10 (41.7%) | 0 | - |
| Genitourinary cancer<sup>c</sup> | 4 (16.7%) | 0 | - |
| Melanoma | 4 (16.7%) | 7 (70%) | - |
| Gastrointestinal cancer<sup>d</sup> | 4 (16.7%) | 2 (20%) | - |
| Head and neck cancer | 2 (8.3%) |  | - |
| Lymphoma | 0 | 1 (10%) | - |
| ICI regimen (N, %) |  |  |  |
| Anti-PD-1 therapy | 20 (83.3%) | 10 (100%) | - |
| Anti-PD-L1 therapy | 2 (8.3%) | - | - |
| Anti-CTLA-4/PD-1 combination | 2 (8.3%) | - |  |
| Concurrent chemotherapy | 4 (16.7%) | 2 (20%) | - |
| Cumulative ICI doses prior to sampling (cycles, IQR) | 6 (4, 8)<sup>*</sup> | 8.5 (4, 18) | - |
| Time from last ICI dose to sIL-2R collection (days, IQR) | 21 (19.5, 29) | 25.5 (20, 56) | - |
| Concurrent irAEs (N, %) | 6 (25.0%) | 0 (%) | - |
| Cancer response to immunotherapy |  |  |  |
| Stable Disease | 21 (87.5%) | 9 (90%) | - |
| Progressive Disease | 3 (12.5%) | 1 (10%) | - |

**Figure 1.**
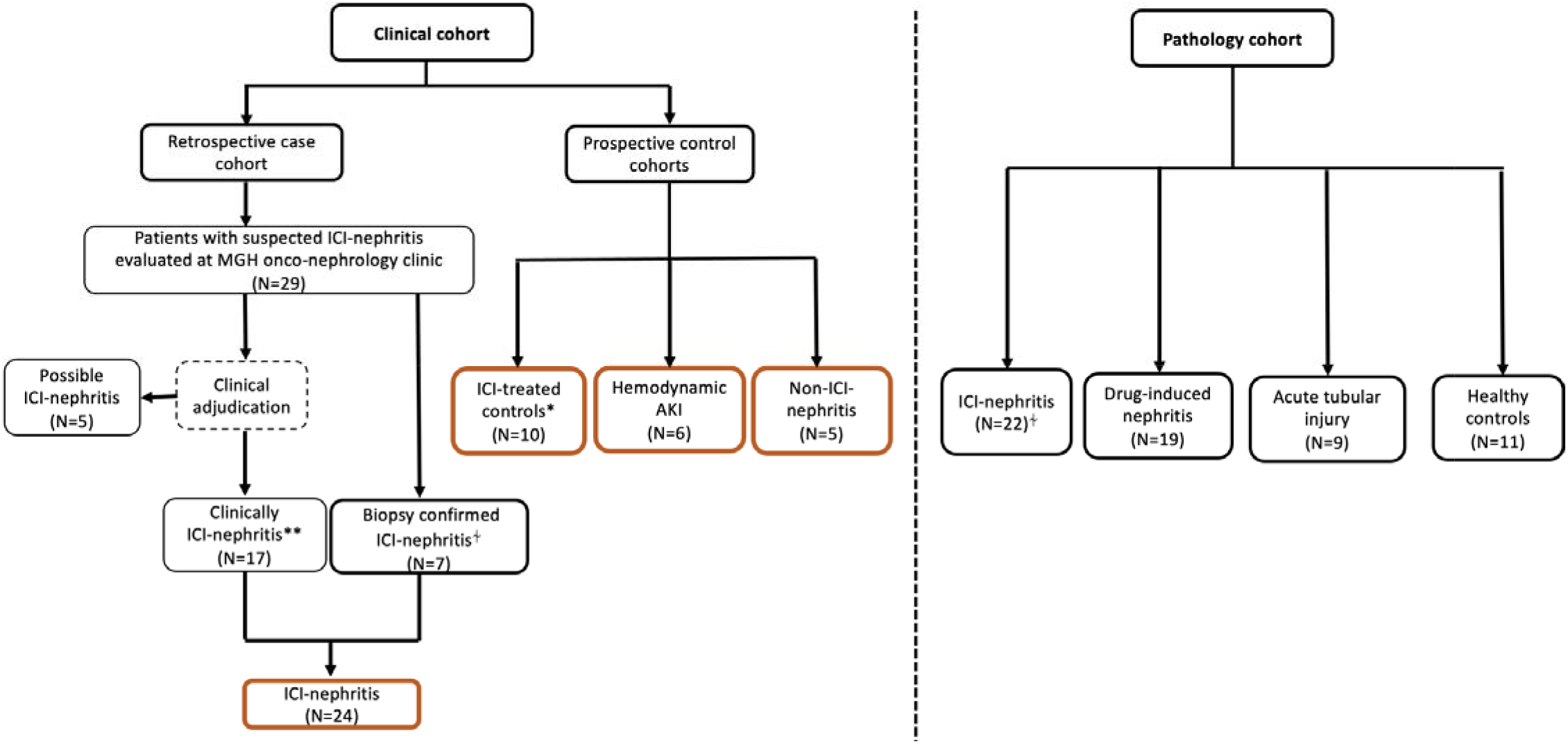
Flow diagram illustrating cohort inclusion. ICI-nephritis, ICI-treated controls, and hemodynamic AKI controls were used to generate ROC for diagnostic performance of sIL-2R in ICI-nephritis; ICI-nephritis and ICI-treated controls were compared to generate ROC for diagnostic performance of peripheral T and B cell marker changes in ICI-nephritis. The four pathology cohorts were compared in nanostring analysis using historical data. Four out of the 7 patients from the clinical cohort who underwent kidney biopsy were also included in the pathology cohort. *ICI-treated controls had normal kidney function and no history of immune related adverse events at the time of inclusion. **Clinically ICI-nephritis requires agreement of the diagnosis between two nephrologists during independent chart review.

### Soluble IL-2 receptor

sIL-2R level in peripheral blood was significantly higher in patients with ICI-nephritis (median 2.5-fold-ULN, IQR 1.9-3.3) as compared to ICI-treated controls (median 0.8-fold-ULN, IQR 0.5-0.9, *P*<0.001) and patients with hemodynamic AKI (median 0.9-fold-ULN, IQR 0.7-1.1, *P*<0.001). There was a wide range of values in the group with possible ICI-nephritis (median 1.6-fold-ULN, IQR 1.2-2.1) in line with the potential diagnostic heterogeneity of this group. Of note, patients with non-ICI-nephritis (all biopsy-proven) had an elevated sIL-2R level comparable to that of ICI-nephritis (median 2.7-fold-ULN, IQR 1.5-3.8, *P*=0.76) (**Figure 2A**).

**Figure 2.**
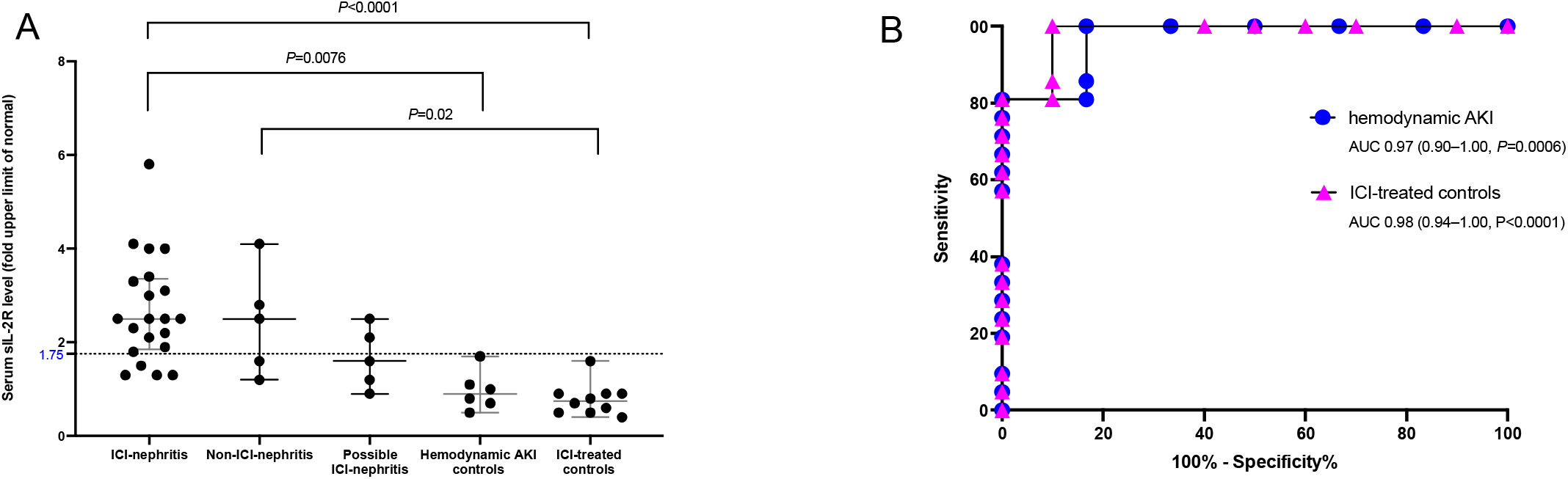
sIL-2R levels and diagnostic performance in patients with ICI-nephritis. **Figure 2A**. Peripheral blood sIL-2R level, shown as fold ULN for each patient group as indicated. Dotted line indicates the optimal cut-off point determined by receiving operating characteristic (ROC) curve analysis (1.75-fold ULN, **Fig. 2B**). Symbols represent unique individuals, bars represent medians (+/-IQRs) of total indicated patients. ANOVA test with significant *P* values shown. **Figure 2B**. Receiver operating characteristic (ROC) curve, comparing ICI-nephritis cases and ICI-treated controls (pink triangles) or hemodynamic AKI controls (blue circles), respectively. Area under the curve (AUC), 95% confidence interval (CI), and *P* value shown.

A receiver operating characteristic (ROC) curve was performed, comparing confirmed ICI-nephritis to ICI-treated controls and confirmed ICI-nephritis to hemodynamic AKI. A sIL-2R cut-off point of 1.75-fold ULN yielded a specificity of 100% (95% CI, 61%-100%) with a sensitivity of 81% (95% CI, 60%-92%) when using hemodynamic AKI as a comparator group, and a specificity of 100% (95% CI, 72%-100%) with a sensitivity of 81% (95% CI, 60%--92%) when using ICI-treated controls as comparator group (**Figure 2B**). Area under the curve (AUC) was >0.96 when using either comparator group as a control.

### Flow cytometry

Given the utility of peripheral blood flow cytometry in the diagnosis of congenital CTLA4 deficiency (16, 17), we compared CLIA-certified flow cytometry panels among patients with ICI-nephritis (N=19) and ICI-treated controls (N=6) who had flow cytometry performed. In patients with ICI-nephritis, we observed lower circulating counts of total CD3+ T cells (geometric mean 583.8 vs. 1333 cells/uL, *P*=0.014), including significantly lower cytotoxic CD8+ T cells (geometric mean 138.6 vs. 444.1 cells/uL, *P*=0.0019) and a trend towards lower helper CD4+ T cells (geometric mean 404.5 vs. 759.2 cells/uL, *P*=0.08) (**Figure 3A**). As loss of naïve T cells in peripheral blood is a prominent feature of CTLA4 deficiency, we further investigated populations of naïve (CD45RA+) and memory (CD45RO+) T cells in circulation. We observed lower absolute counts of naïve cytotoxic CD45RA+CD8+ T cells in patients with ICI-nephritis compared to ICI-treated controls (**Figure 3B**). As individual diagnostic parameters, lower peripheral blood total CD8+ T cells and naïve CD45RA+CD8+ T cells had excellent AUCs for the diagnosis of ICI-nephritis among patients receiving ICI (total CD8+ T cells: AUC=0.90; 95% CI 0.78-1.03; *P*=0.0034 and naïve CD45RA+CD8+ T cells: AUC=0.82; 95% CI 0.66-0.99; *P*=0.019). The likelihood ratio for ICI-nephritis was 4.7 in individuals with a circulating CD8+ T cell level <268 cells/uL and 4.4 in individuals with a circulating naïve CD45RA+CD8+ T cell level <122 cells/uL (**Figure 3C**). Low absolute counts of CD8+ T cells and naïve CD45RA+CD8+ T cells in circulation were observed to be overlapping immunophenotypes with a positive and significant linear correlation identified (R^2^=0.77; *P*<0.0001; **Figure 3D**).

**Figure 3.**
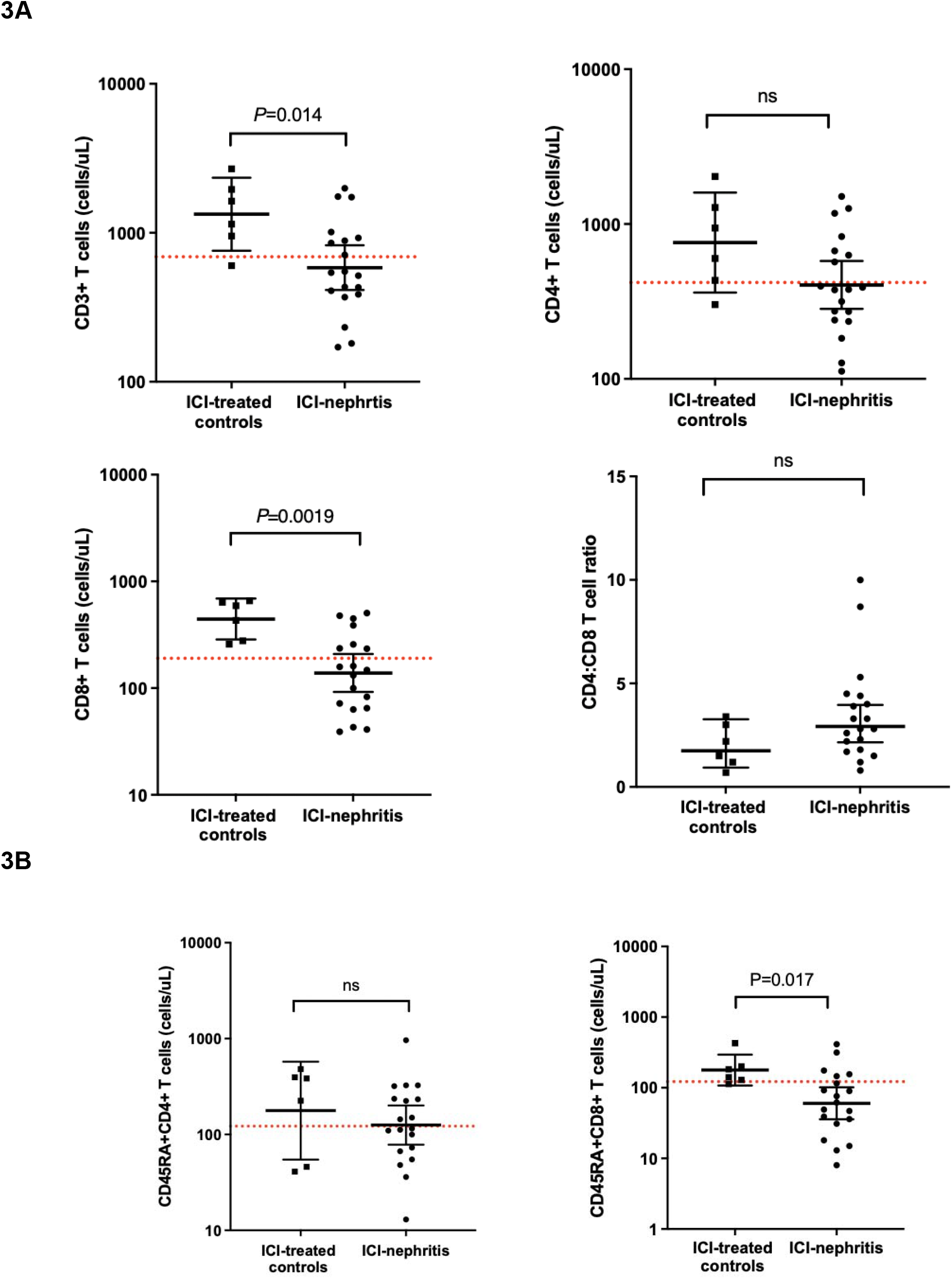
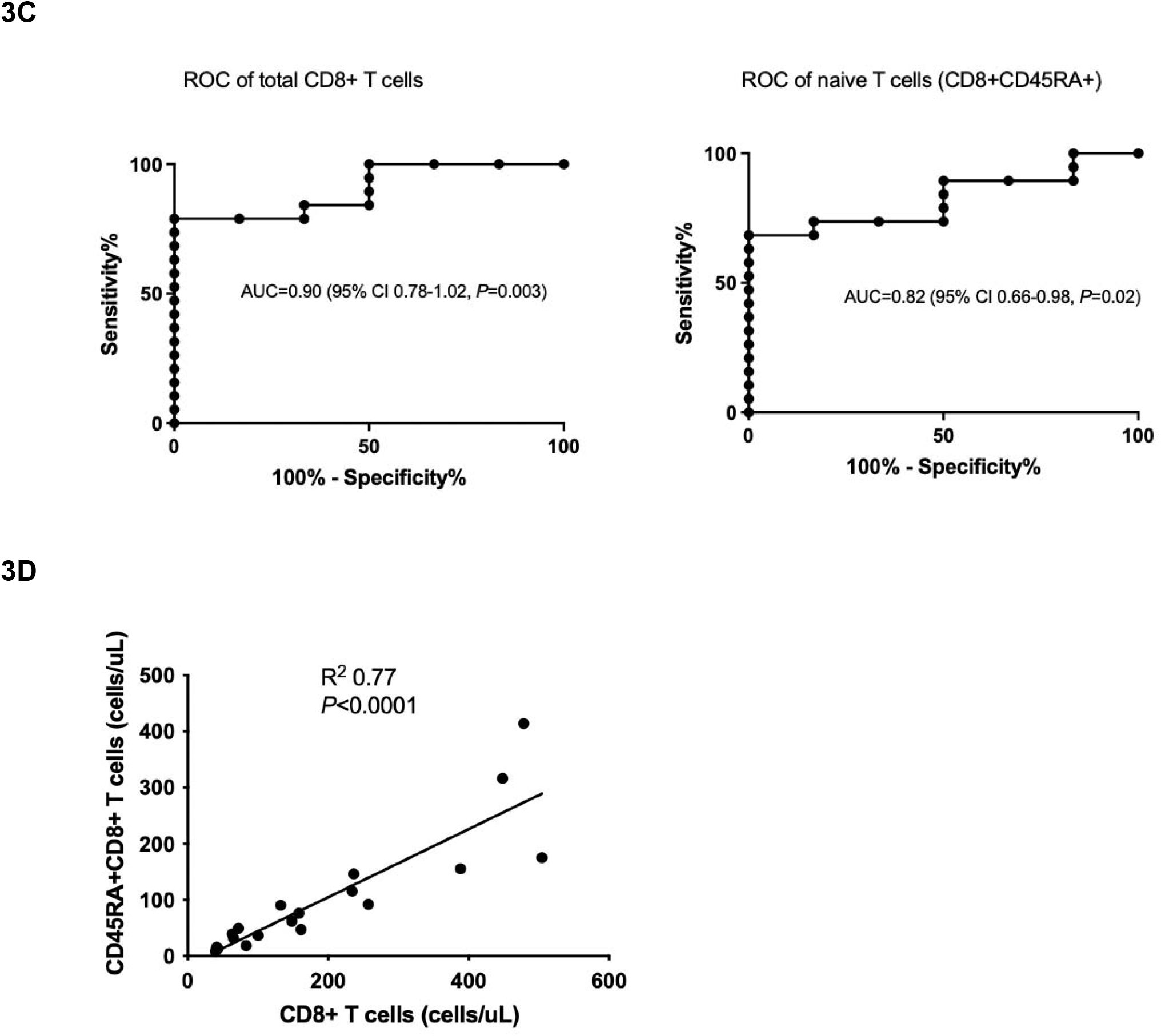
Peripheral blood T cell markers are altered in ICI-nephritis. **(A, B)** T cell subsets, shown as absolute count in log scale or CD4+/CD8+ ratio and compared between ICI-treated controls (N=6; squares) and ICI-nephritis (N=19; circles). Symbols represent unique individuals; bars represent geometric means (95% confidence intervals) of total indicated patients. Red dotted lines indicate the lower limit of normal of the assay. ns=non-significant. **(C)** ROC curves of total CD8+ T cells (left) and naïve CD45RA+CD8+ T cells (right). Area under the curve (AUC), 95% confidence interval (CI), and *P* values shown. **(D)** Linear correlation between total CD8+ T cell count and CD45RA+CD8+ T cell count. R^2^ and P value are shown in the graph. Symbols represent unique individuals; straight line represents fitted regression line.

Patients with CTLA4 deficiency additionally demonstrate prominent B cell defects in peripheral blood that include increased B cell activation and atypical B cell memory development (16, 17). Analysis of total circulating CD19+ B cells identified a trend towards out of range, both low and high, B cells in the majority (52.6%) of patients with ICI-nephritis (**Figure 4A**). In addition, one quarter of ICI-nephritis patients demonstrated plasmablast expansion consistent with a trend towards B cell activation in peripheral blood; in contrast, no plasmablast expansion was identified among individuals in the ICI-treated control group (**Figure 4B**). Finally, lower absolute counts of memory (CD27+) B cells were identified in the blood of patients with ICI-nephritis (geometric mean 8.1 vs 22.5 cells/uL, *P*=0.032). As an individual diagnostic parameter, peripheral blood memory CD27+CD19+ B cells had an AUC of 0.80 (95% CI 0.57-1.02; *P*=0.033) for the diagnosis of ICI-nephritis among patients receiving ICI, and the likelihood ratio for ICI-nephritis was 5.0 in individuals with a circulating memory B cell count <20.5 cells/uL (**Figure 4C**). These B cell findings were non-overlapping immunophenotypes with demonstration of poor concordance by linear relationship to each other as well as the T cell phenotypes described (**Supplemental Figure S1**).

**Figure 4.**
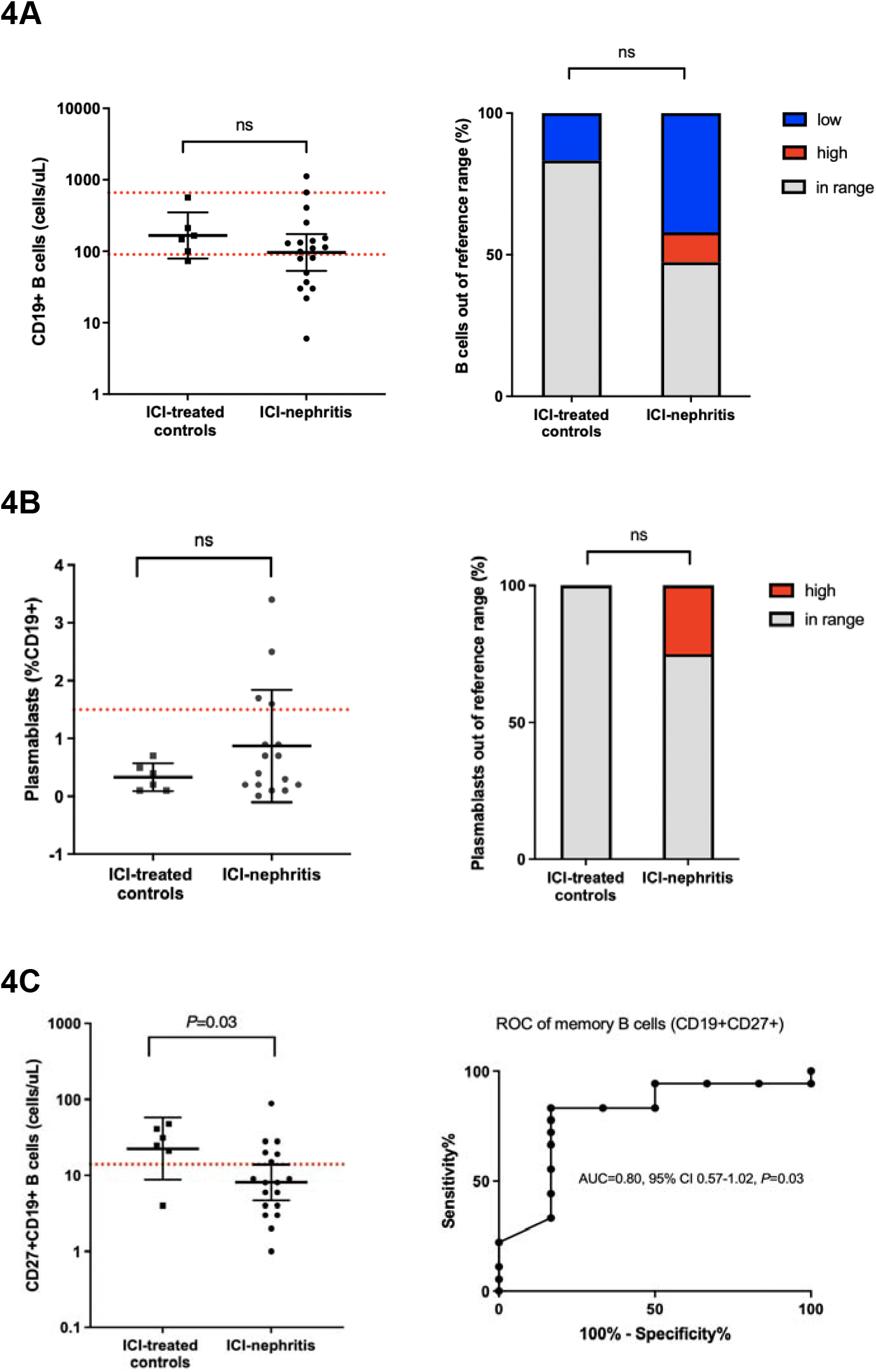
Peripheral blood B cell markers are altered in ICI-nephritis. **(A—C)** B cell subsets, shown as absolute count in log scale, plasmablasts as % total CD19+ B cells, or out-of-range B cells as % total unique individuals as indicated and compared between ICI-treated controls (N=6; squares) and ICI-nephritis (N=19; circles). Symbols represent unique individuals; bars represent geometric means (95% confidence intervals) of total indicated patients. Red dotted lines indicate the assay lower or higher limit of normal. ROC curve of peripheral memory B cells (CD27+CD19+) shown in 4C (right). Area under the curve (AUC), 95% confidence interval (CI), and *P* values shown.

These data suggested a potential benefit from developing a flow cytometry score combining these B cell and T cell components. We created both basic (CD8+ and CD19+), expanded (CD8+, CD19+, CD45RA+CD8+, CD27+CD19+), and complete (CD8+, CD19+, CD45RA+CD8+, CD27+CD19+, and plasmablasts) flow cytometry composite scores for the diagnosis of ICI-nephritis for direct utility in clinical settings given differences between laboratories in flow cytometry capabilities. Using the basic composite score, we found an AUC of 0.84 (95% CI 0.68-1.00, *P*=0.013) in the diagnosis of ICI-nephritis among patients receiving ICIs, which improved further with the expanded and complete composite scores, respectively (**Figure 5**). Overall, scoring by the complete composite score (CD8+, CD19+, CD45RA+CD8+, CD27+CD19+, and plasmablasts) had the highest AUC in the diagnosis of ICI-nephritis among patients receiving ICI (AUC=0.96; 95% CI 0.90-1.03; *P*<0.001). Sensitivity analysis comparing patients with biopsy-confirmed ICI-nephritis to clinically-adjudicated ICI-nephritis did not demonstrate any significant differences in IL-2R or flow cytometry (**Supplemental Figure 2**).

**Figure 5.**
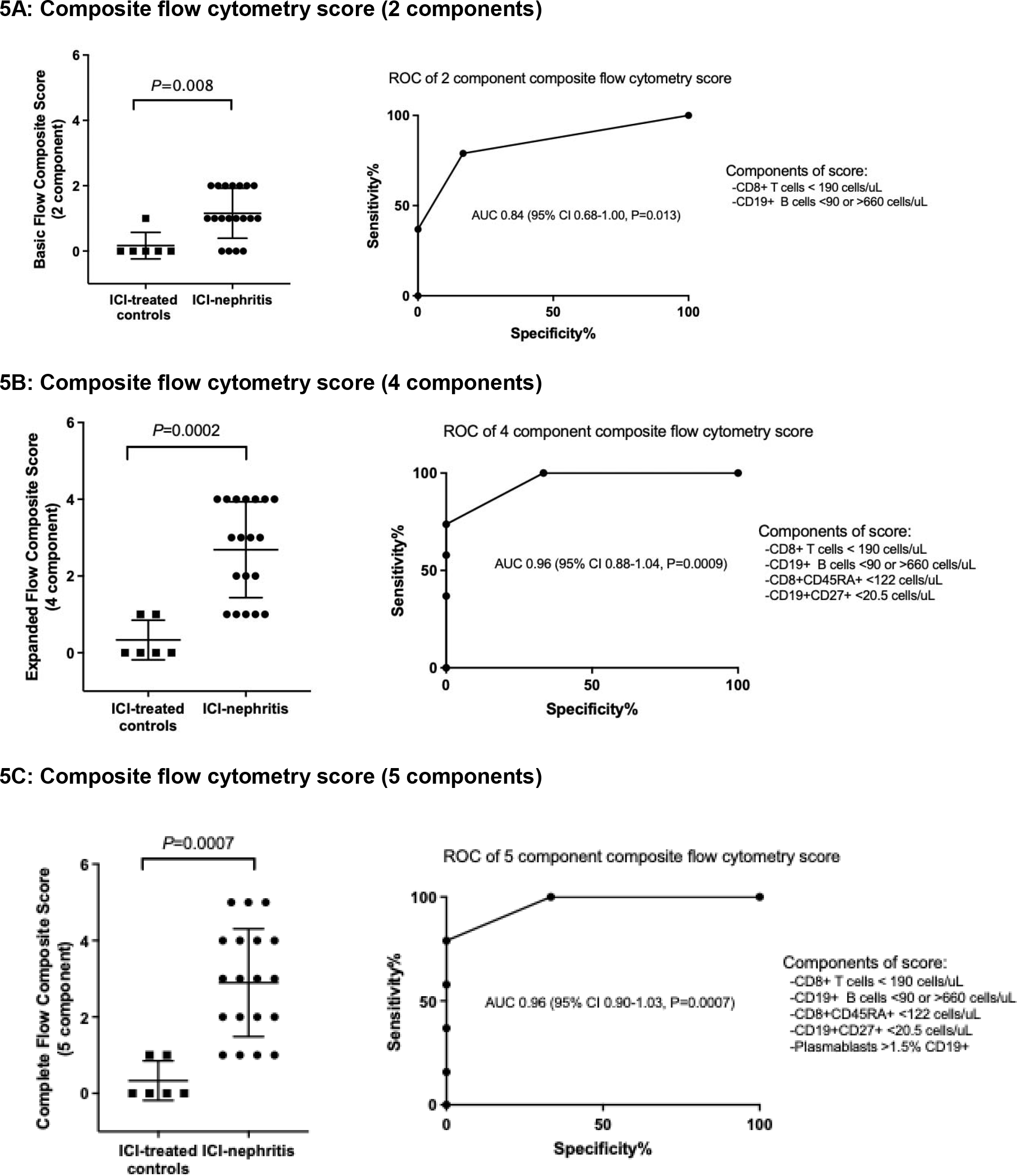
Diagnostic performance of composite B and T cell flow cytometry scores for ICI-nephritis 5A: Composite flow cytometry score (2 components) Diagnostic performance of different composite flow cytometry scores. Scoring is 1 or 0 for each individual component (1 if fulfilling the scoring criteria). Symbols represent unique individuals; bars represent median (95% confidence intervals) of total indicated patients. Area under the curve (AUC), confidence interval (CI), and *P* values shown.

### Kidney biopsy samples

Using archived kidney biopsy samples, we compared transcripts from cases of ICI-nephritis (N=22) to cases without history of ICI treatment: drug-induced nephritis (N=19), acute tubular injury (N=9), and healthy controls (N=11) (**Figure 1**). *IL2RA* gene expression was significantly higher in kidney tissue from cases of ICI-nephritis and drug-induced nephritis, compared to acute tubular injury and controls. IL-2 signaling pathway score and T cell receptor pathway scores were significantly higher in ICI-nephritis and drug-induced nephritis compared to acute tubular injury and controls (**Figure 6**). There were no significant differences in *IL2RA* gene expression, IL-2 signaling pathway score, or T cell receptor pathway scores between ICI-nephritis and other causes of drug-induced nephritis.

**Figure 6.**
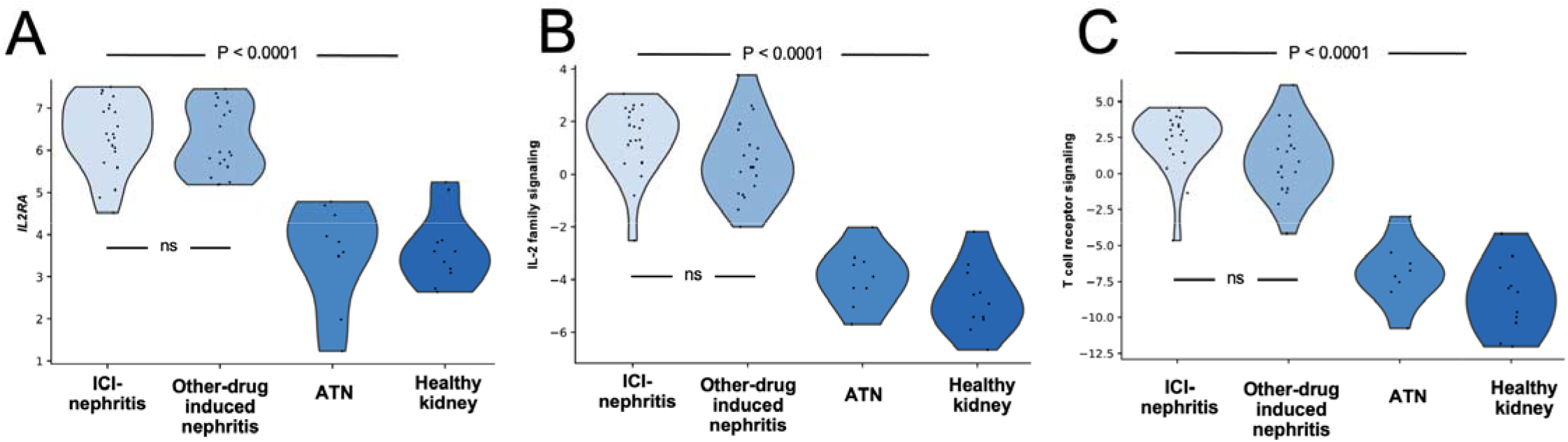
Comparison of *IL2RA* gene expression, T cell receptor scores, and IL2 family signaling in kidney tissue. Gene expression data obtained from FFPE kidney biopsy tissue of patients with ICI-nephritis (N=22), other drug-induced nephritis (N=19), acute tubular necrosis (N=9), and histologically normal control biopsies (N=11). Gene expression shown by violin plot for *IL2RA* in log2 normalized count (**A**). IL-2 family signaling pathway score (**B**) and T cell receptor signaling pathway score (**C**). The pathway score is equal to the first principal component of the gene sets shown in **Supplemental Table 3**. In all cases there was no significant difference between ICI-nephritis and other forms of AIN and the differences between AIN and ATN and healthy control were highly statistically significant. There was no significant difference between ICI-nephritis and other drug-induced nephritis.

## Discussion

We found that patients with ICI-nephritis had soluble and cell-based peripheral blood biomarker abnormalities that mirror those found in patients with CTLA4 deficiency, a congenital T cell hyperactivation syndrome. A sIL-2R cut-off point of 1.75-fold-ULN offered maximal specificity (100%) and optimal sensitivity (81%) in differentiating ICI-nephritis from the ICI-treated controls and hemodynamic AKI controls in this study. We observed that lower absolute counts of circulating CD8+ and naïve CD45RA+CD8+ T cells, as well as lower absolute counts of circulating memory CD27+CD19+ B cells with a relative expansion of plasmablasts in patients with ICI-nephritis compared to ICI-treated controls. A composite flow cytometry score that incorporated T and B cell alterations in peripheral blood, including low total and naïve CD8+ T cells, low CD27+ memory B cells, and a higher percentage of plasmablasts, was shown to have an AUC of 0.96 (95% CI 0.90-1.03; *P*<0.001). These pathologic alterations in peripheral blood flow cytometry mirrored changes seen in patients with congenital CTLA4 deficiency and were not found in patients treated with ICIs who did not have irAEs. Furthermore, we confirmed increased *IL2RA* gene expression in kidney biopsy specimens from patients with ICI-nephritis and other drug-induced interstitial nephritis compared to kidney biopsies from healthy donors or patients with acute tubular injury.

Most of the patients in our study were treated with PD-1/PD-L1 inhibitors not in combination with CTLA4 inhibition (**Table 1**). While CTLA4 signaling occurs prominently in T regulatory cells and PD-1/PD-L1 signaling occurs predominantly in peripheral tissues (4), both are fundamental mechanisms that control peripheral T cell activation and self-tolerance. This may explain why the downstream effect of immunotoxicity on peripheral circulating lymphocytes and inflammatory markers are comparable regardless of the ICI class received and mimic congenital CTLA4 deficiency. Furthermore, abatacept, an immunoglobulin fusion protein that serves as CTLA-4 agonist, has also been successfully used in treatment of severe irAEs induced by either CTLA-4 or PD-1/PD-L1 inhibitors, suggesting overlapping pathogenic pathways of immune-related toxicity shared by all class of ICIs (23).

sIL-2R is the circulating form of the IL-2Rα chain (aka CD25) that was first described in the serum of patients with human T lymphotropic retrovirus type1-associated adult T-cell leukemia (24). IL-2Rα is induced upon activation of T lymphocyte and joins the constitutionally expressed dimeric IL-2R receptor (which contains β and γ chain), and is further enhanced in the subsequent positive feedback loop through IL-2/IL-2R signaling (25). Historically, sIL-2R served as a valuable phenotypic marker for distinguishing adult T-cell leukemia from other lymphocytic neoplasms prior to the wide use of flow cytometry (24, 26). sIL-2R is currently used to support clinical diagnosis of autoimmune disorders characterized by T cell activation, including hemophagocytosis, sarcoidosis, and IgG4 related disease (25, 27), and is generally regarded as a marker of T cell activation. Our finding that most of the ICI-treated controls had sIL-2R levels within the normal range suggests that exposure to ICI alone does not lead to a T cell hyperactivation state sufficient to induce chronic sIL-2R elevation. In fact, the only patient in the ICI-treated control group whose sIL-2R level was above the upper limit of normal had a history of Hodgkin’s lymphoma and was receiving treatment with brentuximab/dacarbazine/ nivolumab at the time of sample collection (**Table 1**). Hematologic malignancies are known to be associated with elevated sIL-2R, potentially decreasing the utility of this biomarker in this specific patient demographic (26). Elevated sIL-2R levels have been reported to associate with tumor burden and disease progression, reflecting higher amount of activated T cells infiltrating the tumor milieu, especially in more immunogenic cancer types such as melanoma and renal cell carcinoma (28-31). Fortunately, in both cohorts of ICI-nephritis and ICI-treated controls, nearly 90% of patients had stable disease at the time of sample collection (**Table 1**). Future research should stratify tumor response and cancer types when evaluating the diagnostic performance of sIL-2R. An increase in sIL-2R level has also been described in patients with chronic kidney disease receiving maintenance hemodialysis by Takamatsu *et al*. (32) and the mechanism is thought to be related to decrease in renal metabolism and/or clearance of sIL-2R; however, our study showed that acute change in kidney function alone does not appear to significantly alter the kinetics of sIL-2R, as patients in the hemodynamic AKI control group did not have elevated sIL-2R levels (**Figure 2A)**.

Previous studies on peripheral blood flow cytometry changes in ICI-treated patients have included B cell markers. A study by Das *et al*. examining patient with melanoma who developed irAEs following ICI described a relative increase in plasmablasts and CD21^lo^ B cells, both of which are associated with a heightened state of B cell activation (33). Moreover, the magnitude of these peripheral blood B cell phenotypic changes directly correlated with the severity of the observed irAEs (33). In our study comparing patients with ICI-nephritis and ICI-treated controls, we found significant changes in peripheral T cell markers, including a decrease in total CD3+ T cells (driven predominantly by lower absolute counts of CD8+ T cells) and loss of naïve circulating CD8+ T cells. In addition, we identified significant changes in peripheral B cell markers, including lower circulating absolute counts of memory CD27+ B cells and a relative expansion of plasmablasts. These changes mirror the peripheral flow cytometry changes observed in patients with congenital CTLA4 deficiency (16).

Robust T cell infiltrates are a classic pathologic feature of irAEs involving different organ systems (34, 35) as well as non-ICI drug-induced acute interstitial nephritis (36). We examined kidney tissue from patients with ICI-nephritis and identified an increase in gene expression for *IL2RA*, IL-2 signaling, and T cell receptor signaling, which mirrored the signature of T cell hyperactivation observed in peripheral blood. Taken together, these finding suggest that activated T cell infiltration of the target organs leads to decline in peripheral T cell populations, which may be central to the pathogenesis of ICI-nephritis. The mechanisms of ICI-nephritis are difficult to study given ICI-nephritis does not occur in animal models(37). Therefore, it is important to study human tissue to further elucidate the immunological mechanisms in ICI-nephritis.

Identifying non-invasive biomarkers of ICI-nephritis is a major unmet need. Traditional biomarkers offer limited accuracy in diagnosis as only one third of patients with ICI-nephritis have leukocyturia, fewer than 15% have white blood cell casts, and “allergic” symptoms such as fever, rash, and eosinophilia are extremely rare (5, 38, 39). Thus, kidney biopsy is often needed for definitive diagnosis, however, biopsy is invasive and is associated with high periprocedural complications in the cancer population (40, 41). The 2021 Society for Immunotherapy of Cancer clinical practice guideline recommended that kidney biopsy should be strongly considered when feasible for patients with stage 2 or 3 AKI (creatinine more than 2 times baseline) (42). However, in real world practice, cohort studies have demonstrated that < 30% of cases of ICI-nephritis are proven by biopsy (43-45). Moledina *et al*. evaluated 32 cases of biopsy-proven acute interstitial nephritis and identified urine tumor necrosis factor alpha and interleukin-9 as biomarkers that significantly improved the diagnostic performance compared to a physician’s clinical assessment. However, this study had very few patients treated with ICIs (N=4), and these findings must be validated in patients receiving ICIs (46). Isik *et al*. compared a cohort of patients with ICI-nephritis (both clinically diagnosed and biopsy proven cases, N=37) to patients with non-ICI-associated AKI (N=13) and found that that serum C-reactive protein (CRP), a systemic inflammatory marker, and urinary retinol binding protein to creatine ratio, a marker for proximal tubular injury, were both significantly elevated at the time of AKI in ICI-nephritis compared to the non-ICI-AKI controls. A normal CRP and urine retinol binding protein to creatinine ratio appear to have a high negative predictive value for ruling out ICI-associated AKI, however, the positive predictive value of elevated biomarkers may be limited (44). Prior studies that evaluated aggregate cytokine scores to diagnose non-kidney irAEs did not include sIL-2R (47). CLIA-certified sIL-2R assays are performed on serum samples only and cannot be performed on frozen plasma, which may explain why it may have been excluded from prior studies. Both sIL-2R and peripheral blood flow cytometry are clinically available tests and flow cytometry, in particular, has a rapid turn-around (less than 24 hours at our center).

Our study has several limitations. First, the size of our control groups are limited, and we did not have a control group of ICI-treated patients who developed AKI due to causes other than ICI-nephritis given the challenges of case identification. Second, we excluded patients experiencing extra-renal irAEs to isolate the effect of ICI-nephritis on sIL-2R and lymphocyte markers. However, given that it is not an organ specific marker, sIL-2R may also be elevated in patients experiencing extra-renal irAEs (48, 49). Prior and/or concomitant extra-renal irAEs is a frequent finding and occurs in nearly half of the patients with ICI-nephritis (38, 39), and thus the diagnostic utility of sIL-2R in the real-world setting requires additional research. Third, in line with the biopsy rate in real-world practice (43-45), only 7 out of the 24 ICI-nephritis cases received kidney biopsy in our cohort; however, no significant difference in sIL-2R level and peripheral T and B cell populations were found between the biopsy-proven cases and clinically-adjudicated cases (**Supplemental Figure S2**). Finally, twenty percent of the ICI-nephritis and ICI-treated controls in our cohort also received concurrent chemotherapy, which could have affected absolute lymphocyte count by causing bone marrow suppression; however, no significant difference in the peripheral T and B cell populations was found between those who received concurrent chemotherapy and those who received ICI therapy alone (**Supplemental Figure S3**).

In conclusion, we report statistically significant differences in sIL-2R level and dysregulated B and T cell phenotypes detected by flow cytometry in patients with ICI-nephritis compared to controls, findings that mirror the immunophenotype found in patients with congenital CTLA4 deficiency. Validating these findings in larger prospectively collected populations of ICI-treated patients with AKI, other irAEs that occur after ICIs, and expanding to other forms of acute interstitial nephritis will be the critical next steps for future research.

## Data Availability

All data produced in the present study are available upon reasonable request to the authors

## Disclosure

RJS receives grant support from Merck, serves as a consultant for BMS, Eisai, Iovance, Merck, Novartis, Pfizer and OncoSec; MES serves as a consultant for Mallinckrodt; JF receives grant support from Pfizer and Bristol Myers Squibb. QW, HS, DM, DH, RNS, IAR, RBC, SC, RF, NY, LC, ACV, KR do not have conflict of interest to declare.

## Funding

MES is funded by NIH R01DK130839. ACV is funded by NIH DP2CAS247831. SB is supported by NIH K23AI163350. SH is funded by NIH K08DK118120 and Mayo CCaTS grant number UL1TR002377.

---

**Supplemental Figure 1.**
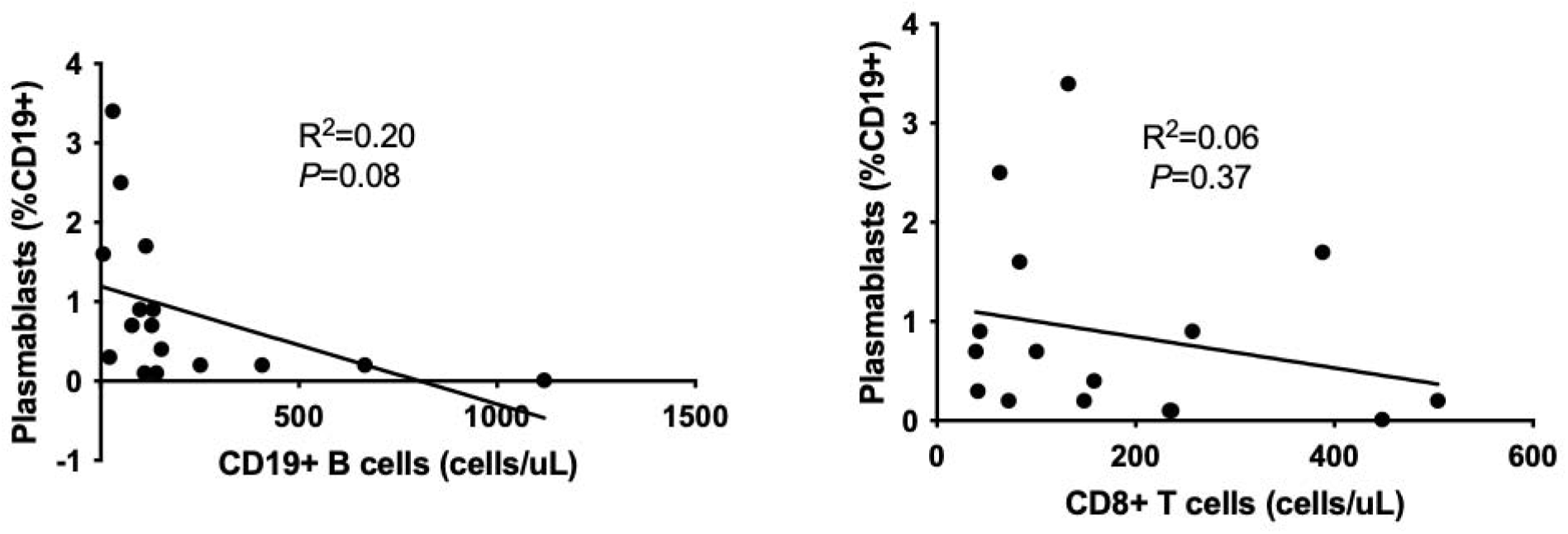
Linear correlation between B cell phenotypes and CD8+ T cells and within different B cell phenotypes. Linear correlation between percentage of plasmablasts and total CD19+ B cell count (left), percentage of plasmablast and CD8+ T cells (right). Symbols represent unique individuals; straight line represents fitted regression line; R^2^ and P values are indicated in the graphs.

**Supplemental Figure 2.**
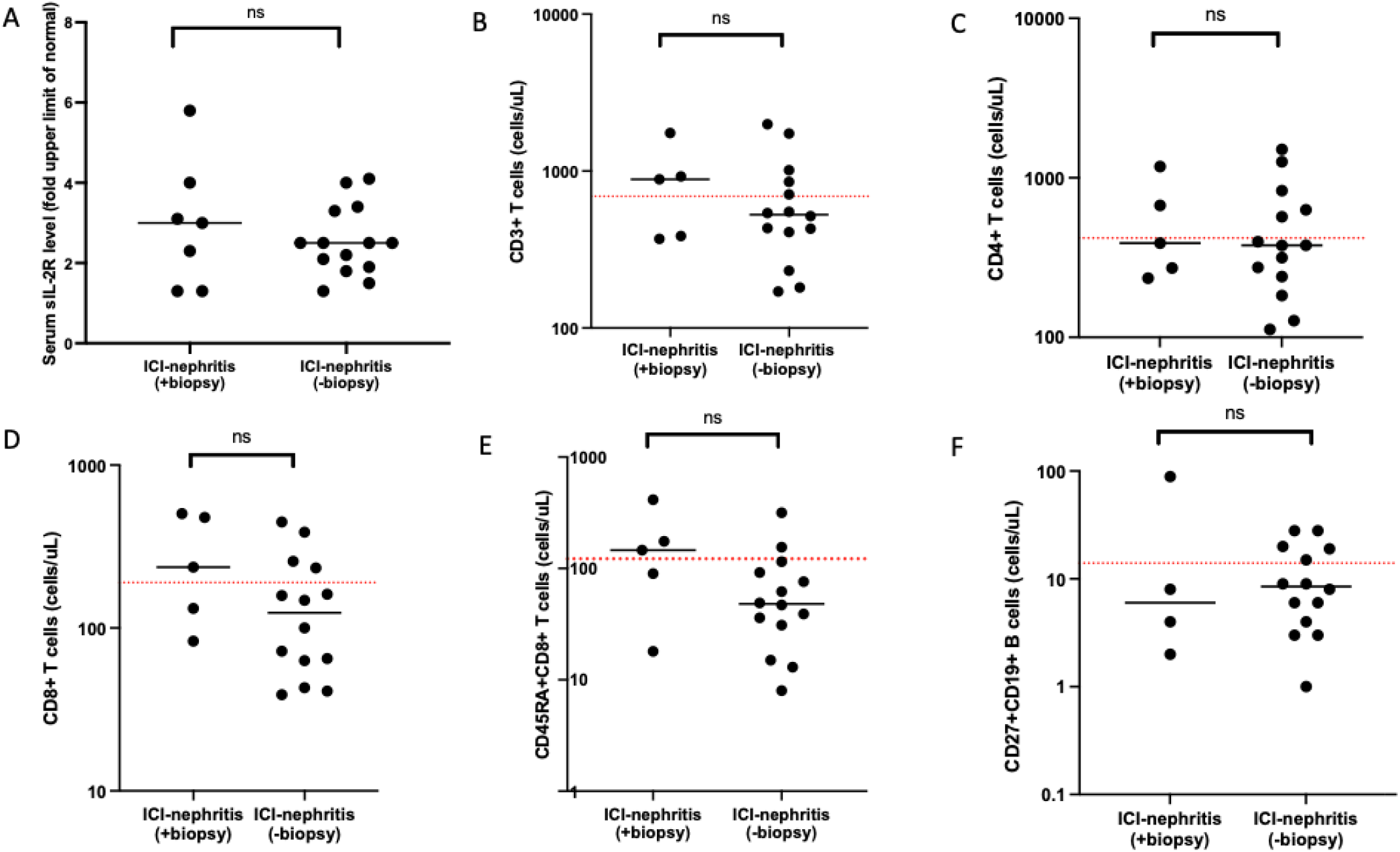
Comparison of sIL-2R level, peripheral T and B cell markers in patients with ICI-nephritis who were diagnosed with kidney biopsy and who were diagnosed by clinical adjudication. (A) Fold ULN of serum sIL-2R level were compared between patients with ICI-nephritis (N=7) who were diagnosed with kidney biopsy and who were diagnosed by clinical criteria (N=14). (B—F) Absolute total lymphocyte counts as indicated (cells/uL), shown in log scale, were compared between patients with ICI-nephritis who were diagnosed with kidney biopsy (N=5) and who were diagnosed by clinical criteria (N=14) who received concurrent chemotherapy (N=4). Symbols represent unique individuals; bars represent geometric means (95% confidence intervals) of total indicated patients; red dotted line represent lower limit of normal of the assay. ns=non-significant

**Supplemental Figure 3.**
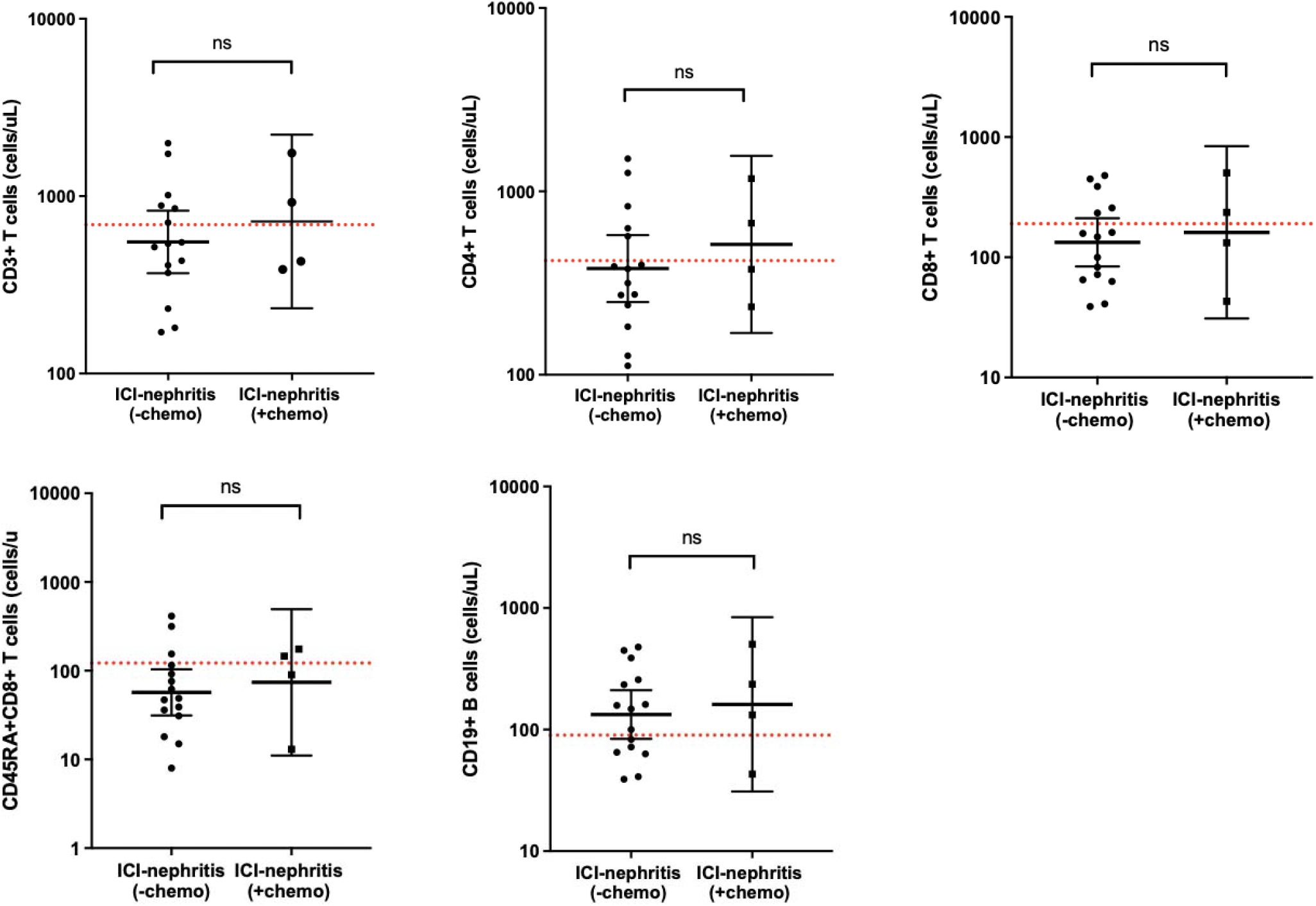
Comparison of peripheral T and B cell markers in patients with ICI-nephritis who received concurrent chemotherapy with ICI or who received ICI therapy alone. Absolute total lymphocyte counts as indicated (cells/uL), shown in log scale, compared between patients with ICI-nephritis who were treated with ICI alone (N=15) and who received concurrent chemotherapy (N=4). Symbols represent unique individuals; bars represent geometric means (95% confidence intervals) of total indicated patients; red dotted line represent lower limit of normal of the assay. ns=non-significant.

**Supplemental Table 1.**
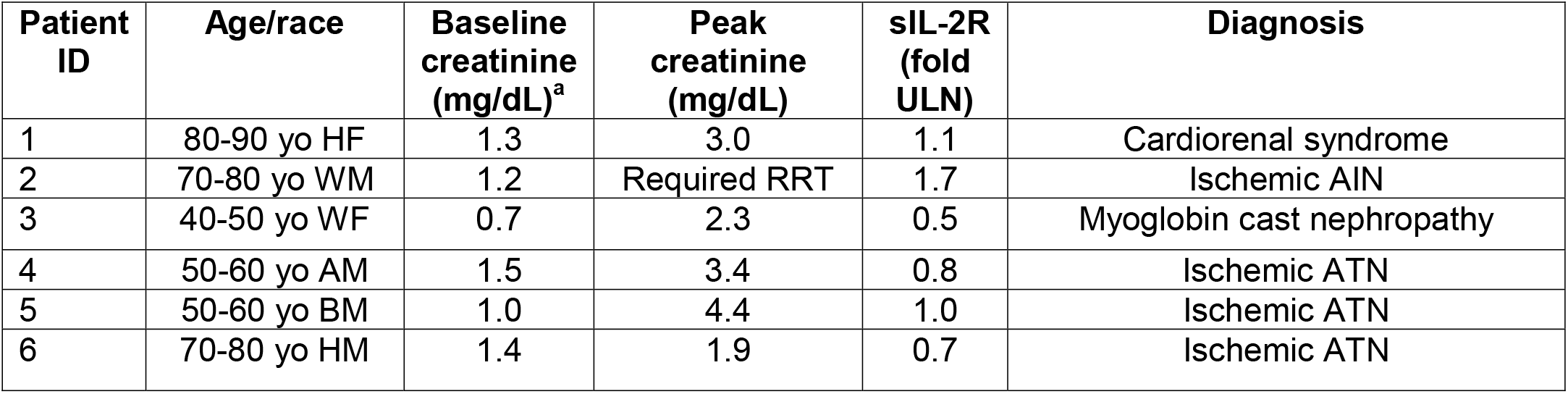
Case summaries for hemodynamic AKI control group. Case summaries for hemodynamic AKI control group. a. case 3,4 and 6 do not have pre-AKI creatinine available within 90 days. Their baseline creatinine was imputed from creatinine nadir during hospitalization. Age ranges in 10-year intervals are used to protect patients’ confidentiality. Abbreviations: sIL-2R=soluble interleukin 2 receptor; HF=Hispanic female; WM=White male; WF=White female; AM=Asian male; BM=Black male; HM=Hispanic male; RRT=renal replacement therapy; AKI=acute kidney injury; ATN=acute tubular necrosis; CKD=chronic kidney disease

**Supplemental Table 2.** Case summaries for non-ICI AIN group. Case summaries for non-ICI-nephritis group. a. case 2 does not have pre-AKI baseline creatinine available within 90 days. Baseline creatine was imputed from creatinine nadir during follow-up course. Age ranges in 10-year intervals are used to protect patients’ confidentiality. Abbreviations: sIL-2R=soluble interleukin 2 receptor; WF=White female; AM=Asian male; BM=Black male; TINU=tubulointerstitial nephritis and uveitis syndrome; PPI=proton pump inhibitor; AIN=acute interstitial nephritis

| Patient ID | Age/race | Baseline creatinine (mg/dL) | Peak creatinine (mg/dL) | sIL-2R (fold ULN) | Diagnosis |
| --- | --- | --- | --- | --- | --- |
| 1 | 50-60 yo WF | 0.8 | 1.6 | 1.2 | TINU syndrome |
| 2 | 70-80 yo AM | 1.5<sup>a</sup> | 5.3 | 4.1 | IgG4-related disease |
| 3 | 30-40 yo WF | 0.9 | 2.7 | 2.5 | TINU syndrome |
| 4 | 30-40 yo WF | 0.9 | 1.1 | 2.8 | TINU syndrome |
| 5 | 70-80 yo BM | 3.0 | 4.5 | 1.6 | PPI-associated AIN |

**Supplemental Table 3.**
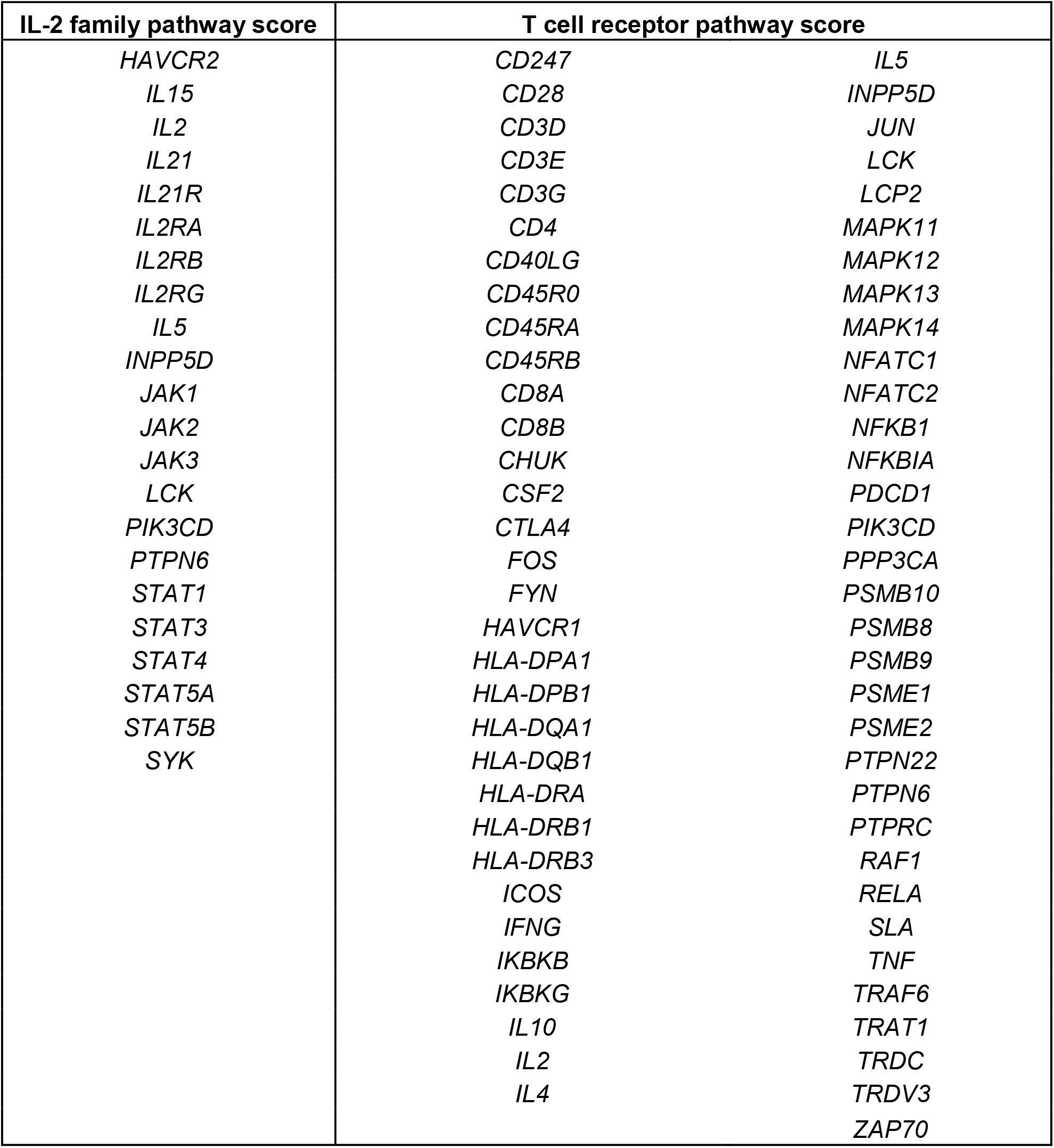
Genes included in Il-2 family pathway score and T cell receptor pathway score. The pathway score is equal to the first principal component of the gene set.

| **IL-2 family pathway score** | **T cell receptor pathway score** |  |
| --- | --- | --- |
| *HAVCR2* | *CD247* | *IL5* |
| *IL15* | *CD28* | *INPP5D* |
| *IL2* | *CD3D* | *JUN* |
| *IL21* | *CD3E* | *LCK* |
| *IL21R* | *CD3G* | *LCP2* |
| *IL2RA* | *CD4* | *MAPK11* |
| *IL2RB* | *CD40LG* | *MAPK12* |
| *IL2RG* | *CD45R0* | *MAPK13* |
| *IL5* | *CD45RA* | *MAPK14* |
| *INPP5D* | *CD45RB* | *NFATC1* |
| *JAK1* | *CD8A* | *NFATC2* |
| *JAK2* | *CD8B* | *NFKB1* |
| *JAK3* | *CHUK* | *NFKBIA* |
| *LCK* | *CSF2* | *PDCD1* |
| *PIK3CD* | *CTLA4* | *PIK3CD* |
| *PTPN6* | *FOS* | *PPP3CA* |
| *STAT1* | *FYN* | *PSMB10* |
| *STAT3* | *HAVCR1* | *PSMB8* |
| *STAT4* | *HLA-DPA1* | *PSMB9* |
| *STAT5A* | *HLA-DPB1* | *PSME1* |
| *STAT5B* | *HLA-DQA1* | *PSME2* |
| *SYK* | *HLA-DQB1* | *PTPN22* |
|  | *HLA-DRA* | *PTPN6* |
|  | *HLA-DRB1* | *PTPRC* |
|  | *HLA-DRB3* | *RAF1* |
|  | *ICOS* | *RELA* |
|  | *IFNG* | *SLA* |
|  | *IKBKB* | *TNF* |
|  | *IKBKG* | *TRAF6* |
|  | *IL10* | *TRAT1* |
|  | *IL2* | *TRDC* |
|  | *IL4* | *TRDV3* |
|  |  | *ZAP70* |

## Notes

### Funding Statement

This study did not receive any funding

### Author Declarations

IRB of Mass General Brigham gave ethical approval for this work

## References

1. Twomey JD, Zhang B. Cancer Immunotherapy Update: FDA-Approved Checkpoint Inhibitors and Companion Diagnostics. The AAPS Journal. 2021;23(2):39.

2. Haslam A, Gill J, Prasad V. Estimation of the Percentage of US Patients With Cancer Who Are Eligible for Immune Checkpoint Inhibitor Drugs. JAMA Netw Open. 2020;3(3):e200423.

3. Ramos-Casals M, Brahmer JR, Callahan MK, Flores-Chávez A, Keegan N, Khamashta MA, et al. Immune-related adverse events of checkpoint inhibitors. Nat Rev Dis Primers. 2020;6(1):38.

4. Perazella MA, Shirali AC. Immune checkpoint inhibitor nephrotoxicity: what do we know and what should we do? Kidney Int. 2020;97(1):62–74.

5. Seethapathy H, Zhao S, Chute DF, Zubiri L, Oppong Y, Strohbehn I, et al. The Incidence, Causes, and Risk Factors of Acute Kidney Injury in Patients Receiving Immune Checkpoint Inhibitors. Clin J Am Soc Nephrol. 2019;14(12):1692–700.

6. Cortazar FB, Marrone KA, Troxell ML, Ralto KM, Hoenig MP, Brahmer JR, et al. Clinicopathological features of acute kidney injury associated with immune checkpoint inhibitors. Kidney Int. 2016;90(3):638–47.

7. Seethapathy H, Zhao S, Strohbehn IA, Lee M, Chute DF, Bates H, et al. Incidence and Clinical Features of Immune-Related Acute Kidney Injury in Patients Receiving Programmed Cell Death Ligand-1 Inhibitors. Kidney Int Rep. 2020;5(10):1700–5.

8. Manohar S, Ghamrawi R, Chengappa M, Goksu BNB, Kottschade L, Finnes H, et al. Acute Interstitial Nephritis and Checkpoint Inhibitor Therapy. Kidney360. 2020;1.

9. Baker ML, Yamamoto Y, Perazella MA, Dizman N, Shirali AC, Hafez N, et al. Mortality after acute kidney injury and acute interstitial nephritis in patients prescribed immune checkpoint inhibitor therapy. J Immunother Cancer. 2022;10(3).

10. Douketis JD, Spyropoulos AC, Spencer FA, Mayr M, Jaffer AK, Eckman MH, et al. Perioperative management of antithrombotic therapy: Antithrombotic Therapy and Prevention of Thrombosis, 9th ed: American College of Chest Physicians Evidence-Based Clinical Practice Guidelines. Chest. 2012;141(2 Suppl):e326S–e50S.

11. Corapi KM, Chen JL, Balk EM, Gordon CE. Bleeding complications of native kidney biopsy: a systematic review and meta-analysis. Am J Kidney Dis. 2012;60(1):62–73.

12. Hogan JJ, Mocanu M, Berns JS. The Native Kidney Biopsy: Update and Evidence for Best Practice. Clin J Am Soc Nephrol. 2016;11(2):354–62.

13. Perazella MA. Clinical Approach to Diagnosing Acute and Chronic Tubulointerstitial Disease. Adv Chronic Kidney Dis. 2017;24(2):57–63.

14. Muriithi AK, Nasr SH, Leung N. Utility of urine eosinophils in the diagnosis of acute interstitial nephritis. Clin J Am Soc Nephrol. 2013;8(11):1857–62.

15. Fogazzi GB, Ferrari B, Garigali G, Simonini P, Consonni D. Urinary sediment findings in acute interstitial nephritis. Am J Kidney Dis. 2012;60(2):330–2.

16. Kuehn HS, Ouyang W, Lo B, Deenick EK, Niemela JE, Avery DT, et al. Immune dysregulation in human subjects with heterozygous germline mutations in CTLA4. Science. 2014;345(6204):1623–7.

17. Schubert D, Bode C, Kenefeck R, Hou TZ, Wing JB, Kennedy A, et al. Autosomal dominant immune dysregulation syndrome in humans with CTLA4 mutations. Nature Medicine. 2014;20(12):1410–6.

18. Alroqi FJ, Charbonnier LM, Baris S, Kiykim A, Chou J, Platt CD, et al. Exaggerated follicular helper T-cell responses in patients with LRBA deficiency caused by failure of CTLA4-mediated regulation. J Allergy Clin Immunol. 2018;141(3):1050-9.e10.

19. Lo B, Zhang K, Lu W, Zheng L, Zhang Q, Kanellopoulou C, et al. Patients with LRBA deficiency show CTLA4 loss and immune dysregulation responsive to abatacept therapy. Science. 2015;349(6246):436–40.

20. Mengel M, Loupy A, Haas M, Roufosse C, Naesens M, Akalin E, et al. Banff 2019 Meeting Report: Molecular diagnostics in solid organ transplantation-Consensus for the Banff Human Organ Transplant (B-HOT) gene panel and open source multicenter validation. Am J Transplant. 2020;20(9):2305–17.

21. Rosales I, Mahowald G, Tomaszewski K, Hotta K, Iwahara N, Otsuka T, et al. Banff Human Organ Transplant Transcripts Correlate with Renal Allograft Pathology and Outcome: Importance of Capillaritis and Subpathologic Rejection. Journal of the American Society of Nephrology. 2022:ASN.2022040444.

22. Tomfohr J, Lu J, Kepler TB. Pathway level analysis of gene expression using singular value decomposition. BMC Bioinformatics. 2005;6(1):225.

23. Salem J-E, Allenbach Y, Vozy A, Brechot N, Johnson DB, Moslehi JJ, et al. Abatacept for Severe Immune Checkpoint Inhibitor–Associated Myocarditis. New England Journal of Medicine. 2019;380(24):2377–9.

24. Rubin LA, Kurman CC, Fritz ME, Biddison WE, Boutin B, Yarchoan R, et al. Soluble interleukin 2 receptors are released from activated human lymphoid cells in vitro. J Immunol. 1985;135(5):3172–7.

25. Dik WA, Heron M. Clinical significance of soluble interleukin-2 receptor measurement in immune-mediated diseases. Neth J Med. 2020;78(5):220–31.

26. Rubin LA, Nelson DL. The soluble interleukin-2 receptor: biology, function, and clinical application. Ann Intern Med. 1990;113(8):619–27.

27. Handa T, Matsui S, Yoshifuji H, Kodama Y, Yamamoto H, Minamoto S, et al. Serum soluble interleukin-2 receptor as a biomarker in immunoglobulin G4-related disease. Mod Rheumatol. 2018;28(5):838–44.

28. Vuoristo MS, Laine S, Huhtala H, Parvinen LM, Hahka-Kemppinen M, Korpela M, et al. Serum adhesion molecules and interleukin-2 receptor as markers of tumour load and prognosis in advanced cutaneous melanoma. Eur J Cancer. 2001;37(13):1629–34.

29. Murakami S. Soluble interleukin-2 receptor in cancer. Front Biosci. 2004;9:3085–90.

30. Bien E, Balcerska A. Serum soluble interleukin 2 receptor alpha in human cancer of adults and children: a review. Biomarkers. 2008;13(1):1–26.

31. Boyano MD, Garcia-Vázquez MD, López-Michelena T, Gardeazabal J, Bilbao J, Cañavate ML, et al. Soluble interleukin-2 receptor, intercellular adhesion molecule-1 and interleukin-10 serum levels in patients with melanoma. Br J Cancer. 2000;83(7):847–52.

32. Takamatsu T, Yasuda N, Ohno T, Kanoh T, Uchino H, Fujisawa A. Soluble interleukin-2 receptors in the serum of patients with chronic renal failure. Tohoku J Exp Med. 1988;155(4):343–7.

33. Das R, Bar N, Ferreira M, Newman AM, Zhang L, Bailur JK, et al. Early B cell changes predict autoimmunity following combination immune checkpoint blockade. The Journal of Clinical Investigation. 2018;128(2):715–20.

34. Johnson DB, Balko JM, Compton ML, Chalkias S, Gorham J, Xu Y, et al. Fulminant Myocarditis with Combination Immune Checkpoint Blockade. New England Journal of Medicine. 2016;375(18):1749–55.

35. Luoma AM, Suo S, Williams HL, Sharova T, Sullivan K, Manos M, et al. Molecular Pathways of Colon Inflammation Induced by Cancer Immunotherapy. Cell. 2020;182(3):655-71.e22.

36. Spanou Z, Keller M, Britschgi M, Yawalkar N, Fehr T, Neuweiler J, et al. Involvement of Drug-Specific T Cells in Acute Drug-Induced Interstitial Nephritis. Journal of the American Society of Nephrology. 2006;17(10):2919–27.

37. Curran MA, Montalvo W, Yagita H, Allison JP. PD-1 and CTLA-4 combination blockade expands infiltrating T cells and reduces regulatory T and myeloid cells within B16 melanoma tumors. Proc Natl Acad Sci U S A. 2010;107(9):4275–80.

38. Cortazar FB, Kibbelaar ZA, Glezerman IG, Abudayyeh A, Mamlouk O, Motwani SS, et al. Clinical Features and Outcomes of Immune Checkpoint Inhibitor-Associated AKI: A Multicenter Study. J Am Soc Nephrol. 2020;31(2):435–46.

39. Gupta S, Short SA, Sise ME, Prosek JM, Madhavan SM, Soler MJ, et al. Acute kidney injury in patients treated with immune checkpoint inhibitors. Journal for immunotherapy of cancer. 2021;9(10):e003467.

40. Moledina DG, Luciano RL, Kukova L, Chan L, Saha A, Nadkarni G, et al. Kidney Biopsy-Related Complications in Hospitalized Patients with Acute Kidney Disease. Clin J Am Soc Nephrol. 2018;13(11):1633–40.

41. Korbet SM, Gashti CN, Evans JK, Whittier WL. Risk of percutaneous renal biopsy of native kidneys in the evaluation of acute kidney injury. Clin Kidney J. 2018;11(5):610–5.

42. Brahmer JR, Abu-Sbeih H, Ascierto PA, Brufsky J, Cappelli LC, Cortazar FB, et al. Society for Immunotherapy of Cancer (SITC) clinical practice guideline on immune checkpoint inhibitor-related adverse events. Journal for ImmunoTherapy of Cancer. 2021;9(6):e002435.

43. Stein C, Burtey S, Mancini J, Pelletier M, Sallée M, Brunet P, et al. Acute kidney injury in patients treated with anti-programmed death receptor-1 for advanced melanoma: a real-life study in a single-centre cohort. Nephrol Dial Transplant. 2021;36(9):1664–74.

44. Isik B, Alexander MP, Manohar S, Vaughan L, Kottschade L, Markovic S, et al. Biomarkers, Clinical Features, and Rechallenge for Immune Checkpoint Inhibitor Renal Immune-Related Adverse Events. Kidney Int Rep. 2021;6(4):1022–31.

45. Oleas D, Bolufer M, Agraz I, Felip E, Muñoz E, Gabaldón A, et al. Acute interstitial nephritis associated with immune checkpoint inhibitors: a single-centre experience. Clin Kidney J. 2021;14(5):1364–70.

46. Moledina DG, Wilson FP, Pober JS, Perazella MA, Singh N, Luciano RL, et al. Urine TNF-α and IL-9 for clinical diagnosis of acute interstitial nephritis. JCI Insight. 2019;4(10).

47. Lim SY, Lee JH, Gide TN, Menzies AM, Guminski A, Carlino MS, et al. Circulating Cytokines Predict Immune-Related Toxicity in Melanoma Patients Receiving Anti-PD-1-Based Immunotherapy. Clin Cancer Res. 2019;25(5):1557–63.

48. Takai R, Funakoshi Y, Suto H, Nagatani Y, Imamura Y, Toyoda M, et al. Serum Soluble Interleukin-2 Receptor as a Potential Biomarker for Immune-related Adverse Events. Anticancer Research. 2021;41(2):1021–6.

49. Yoshida K, Morishima Y, Shiozawa T, Nakazawa K, Matsuyama M, Kiwamoto T, et al. Serum Soluble Interleukin-2 Receptor as a Possible Biomarker for the Early Detection and Follow-up of Nivolumab-Induced Pneumonitis. Journal of Thoracic Oncology. 2019;14(5):e90–e1.

